# Knowledge, attitudes, and positions of religious leaders towards female genital mutilation: A cross-sectional study from the Kurdistan Region of Iraq

**DOI:** 10.1101/2022.03.09.22272143

**Authors:** Kazhan I. Mahmood, Sherzad A. Shabu, Karwan M-Amen, Abubakir M. Saleh, Hamdia Ahmed, Barzhang Q. Mzori, Nazar P. Shabila

**Affiliations:** College of Nursing, Hawler Medical University, Erbil, Iraq; Department of Community Medicine, College of Medicine, Hawler Medical University, Erbil, Iraq; College of Health Sciences, Hawler Medical University, Erbil, Iraq; Erbil Directorate of Health, Erbil, Iraq

**Keywords:** Female genital cutting, Religious leaders, Knowledge, Attitudes

## Abstract

**Background:** Understanding the perspectives of the key players in the community regarding female genital cutting (FGC) is very important for directing preventive programs. Religious leaders help shape community behaviors, which is highly pertinent in the case of FGC as it is frequently perceived to be a religious requirement. This study assesses religious leaders’ knowledge, attitudes, and positions towards FGC in the Kurdistan Region of Iraq.

**Methods:** This cross-sectional study was conducted in the Kurdistan Region of Iraq. It included a purposive sample of 147 local religious leaders (*khateebs*) representing the three governorates of Erbil, Sulaimaniyah, and Duhok. A self-administered questionnaire was used to collect data about the religious leaders’ knowledge, attitude, and position towards FGC.

**Results:** The participants identified reduction of the sexual desire of women as the main benefit (37%) and risk (24%) of FGC. Cultural tradition and religious requirements were the main reasons for practicing FGC. About 59% of the religious leaders stated that people ask for their advice on FGC. Around 14% of the participants supported performing FGC, compared to 39.1% who opposed it. Religious (73.9%) and cultural (26.1%) rationales were the main reasons given for supporting FGC. Being a cultural practice and having harmful effects (53.5%) and lack of clear religious evidence (46.6%) were the main reasons for being against FGC. Around 52% of the participants recommended banning FGC by law, while 43.5% did not support banning it. A statistically significant association (P=0.015) was found between religious leaders’ residence and their position on performing FGC. More than 46% of those residing in Duhok were against performing FGC, compared to lower proportions in Erbil (38.8%) and Sulaimaniyah (30%).

**Conclusion:** A conclusive decision concerning FGC banning needs to be made by religious authorities to advise people to avoid the practice. Health awareness activities incorporating FGC risks should be carried out to enlighten religious leaders at different levels of religious positions. Further research exploring perspectives of religious authorities concerning religious leaders’ inconclusive judgments about FGC is deemed necessary.

## Introduction

Female genital cutting (FGC) involves partial or complete removal of the female external genitalia or other injuries to the genital organ for non-therapeutic reasons [1]. While FGC has no health benefits, it can cause severe bleeding and urinating problems as immediate health consequences. Cysts, infections, childbirth complications, and increased risk of newborn deaths are long-term health consequences [2,3]. More than 200 million girls and women in 30 countries in Africa, the Middle East, and Asia have experienced FGC [4]. FGC is predominantly performed on young girls between infancy and the age of 15 years [5]. The Iraq Multiple Indicator Cluster Survey 2018 showed that 7.4% of women aged 15-49 in Iraq underwent FGC. Most cases (37.5%) were from the Kurdistan Region of Iraq (KRI), compared with only 0.4% in central and southern Iraq [6]. The prevalence varies by geographical location among governorates, from 3.1% in Duhok to 45.1% in Sulaymaniyah, and 50.1% in Erbil [7]. The most frequent type of FGC in the KRI is type I (76-99%), involving partial or total removal of the clitoris and/or the prepuce [8,9].

The history and origin of FGC in the KRI is unclear. FGC practice is common in Iraqi and Iranian Kurdish areas [10], but it is rarely practiced in other parts of Iraq or the Kurdish areas in Syria and Turkey. In Iran, FGC is primarily practiced in the governorates of Hormozgan (60%), West Azerbaijan (21%), Kermanshah (18%), and Kurdistan (16%), all of which share borders with KRI [11]. The World Health Organization does not list Turkey and Syria as countries where FGC is traditionally practiced. In areas where it is a longstanding tradition, it is often attributable to social and cultural traditions [8,9,12,13]. It is believed that FGC reduces sexual desire, which is seen as difficult to control and likely to make uncut women susceptible to sexual promiscuity, and thereby prone to dishonor both themselves and their families [14].

Religious requirements are considered an important cause of practicing FGC in many settings, including in the KRI [8,9,15]. Many people believe that religion, particularly Islam, supports FGC [8,9,16]. Most academic literature on the subject usually denies the presence of religious scripts that explicitly prescribe or encourage FGC [16,17]. The Holy Quran and the *hadiths* containing the words, actions, and customs of the Prophet Muhammad (peace be upon him) are the main pillars of Islamic law. There are authenticated (*sahih*), sound or good (*hasan*), and non-authenticated or weak (*daif*) *hadiths*. FGC is not prescribed in the Holy Quran, but it has been mentioned indirectly in some authenticated *hadiths*, and as a religious obligation in some weak *hadiths* [17].

Understanding the perspectives of the key players in the community regarding FGC is very important for directing preventive programs. Religious leaders can help people to shape their behavior [18]. Such effect can be important in the case of FGC as it is frequently perceived as a religious requirement. Nevertheless, the position of Islam toward FGC is basically unclear among the general public, including among those who practice such activities. On the other hand, different cultural backgrounds, traditions, and socioeconomic factors might influence the views of the religious leaders toward FGC. Therefore, understanding the views of religious leaders is vital for their influence on their communities. This study aimed to assess religious leaders’ knowledge, attitudes, and positions towards FGC in the KRI.

## Materials and Methods

This cross-sectional study was conducted in KRI from January to May 2019. In each of the three governorates of KRI religious leaders were approached and invited through the Kurdistan Religious Union. In each governorate, the religious leaders were gathered in one event at either the Union’s office or a mosque. The researchers explained the purpose and the details of the study to the religious leaders and invited them to provide informed consent and complete a self-administered questionnaire. We noticed that there was reluctance to complete the questionnaire and discuss the topic of FGC among many religious leaders, particularly in the Sulaymaniyah governorate. Despite the continuous follow-up through the Kurdistan Religious Union representatives and directly with the religious leaders, only 147 religious leaders out of 500 returned a completed questionnaire.

The self-administered questionnaire was developed in the local language (Kurdish) by the researchers, which included, in addition to questions on the socioeconomic status of the participants, three main categories of questions, mainly in the form of multiple options that the respondents could choose the options they agree with. The first category of questions was about the religious leaders’ knowledge regarding FGC, including purported benefits of the practice, risks associated with it, reasons for practicing it, who performs it, who decides on its performance, and reasons for the different prevalence of FGC in different governorates. The second category of questions was about the religious leaders’ attitudes towards FGC, including whether people ask for their advice on performing FGC, who asks more and when, whether people complain about performing or not performing FGC, and what the role of religious leaders in combating FGC could be. The third category of questions was about the religious leaders’ position towards FGC, including their personal position towards FGC practice, reasons for supporting or being against FGC, whether they recommend banning FGC and why, who needs to undergo and perform FGC, religious views and the existing religious scripts on FGC, and different scriptural texts available in this regard.

The study was approved by the Research Ethics Committee of Hawler Medical University. All participants were informed in advance about the main objectives of this study, and informed consent was obtained from all the participants before they participated. The anonymity of the participants was ensured throughout the study. Microsoft Excel 2010 and SPSS version 20 were used for data summarization and data analysis purposes. Chi-square was used to identify any association between different variables in the study. A P value of ≤ 0.05 was considered to be statistically significant.

## Results

Of 147 religious leaders who participated in this study, about half of them were 40 years old and younger, and had 11-20 years of work experience. The majority (almost two-thirds) of them were from Erbil city and college/institute graduates, as shown in Table 1.

**Table 1.** Participants’ socio-demographic characteristics.

| Variable | Frequency | Percentage |
| --- | --- | --- |
| **Age groups (years)** |  |  |
| ≤ 40 | 73 | (49.7) |
| > 40 | 74 | (50.3) |
| **Work experience (years)** |  |  |
| ≤ 10 | 41 | (27.9) |
| 11-20 | 75 | (51.0) |
| ≥ 21 | 31 | (21.1) |
| **Educational level** |  |  |
| Primary/secondary | 22 | (15.0) |
| College/institute | 100 | (68.0) |
| Higher education | 25 | (17.0) |
| **Governorate** |  |  |
| Erbil | 95 | (64.6) |
| Sulaimaniyah | 22 | (15.0) |
| Duhok | 30 | (20.4) |
| **Total** | **147** | **(100)** |

Regarding the purported benefits of FGC, more than 37% of the participants mentioned that it reduces or regulates the excessive sexual desire of women to avoid sins and social problems. Around 20% of the participants said it enhances the hygiene and cleanliness of women and prevents bad odor, while 16.2% said it helps avoid extra-marital sex. Concerning the risks attached to performing FGC, 24% of the participants mentioned that it reduces or leads to losing sexual desire, 16.6% said it leads to psychological problems, and 13% stated that it is risk-free. Regarding the reason behind practicing FGC, more than 29% stated it is a cultural tradition, followed by 24.1% who said it is a tradition mixed with religion, and 16.09% said it is for reducing the sexual desire of women. About 33% stated that older women are the main performers of FGC, followed by traditional birth attendants (27.9%) and traditional circumcisers (18.7). More than 31% thought that the mother decides to perform FGC for her daughter, followed by both parents (28.3%), and grandmothers (16.4%). More than 54% thought that FGC is not common in the KRI, compared to only 22.14% who thought it is common, and more than 90% thought that the trend of FGC in the KRI is decreasing. Around 28% thought that the difference in the prevalence of FGC in different governorates is attributed to differences in traditional cultures, followed by the difference in people’s education and awareness level (16.9%) and in schools of religious jurisprudence (15.7%). Table 2 shows the details of the knowledge of religious leaders about FGC.

**Table 2.** Knowledge of religious leaders regarding FGC.

| Variable | Frequency | Percentage |
| --- | --- | --- |
| **Benefits of FGC** |  |  |
| Reduce or regulate the excessive sexual desire of women to avoid sins and social problems | 76 | (37.3) |
| Enhancing the hygiene and cleanliness of women and preventing bad odor | 41 | (20.1) |
| Make food from women's hands halal | 7 | (3.4) |
| Help in maintaining the virginity until marriage | 13 | (6.4) |
| Help in avoiding extra-marital sex | 33 | (16.2) |
| Prevents annoying the men by clitoris during sex | 16 | (7.8) |
| Do not know | 18 | (8.8) |
| **Risks attached to FGC** |  |  |
| It is risk free | 37 | (13.1) |
| Reduced or loss of sexual desire | 68 | (24.0) |
| Bleeding | 29 | (10.3) |
| Pain | 31 | (11.0) |
| Infection | 26 | (9.2) |
| Psychological problems | 47 | (16.6) |
| Birth complications | 19 | (6.7) |
| Others | 11 | (3.9) |
| Do not know | 15 | (5.3) |
| **Reasons for practicing FGC** |  |  |
| It is a religious requirement | 28 | (10.7) |
| It is a cultural tradition | 76 | (29.1) |
| It is a tradition mixed with the religion | 63 | (24.1) |
| It enhances the hygiene of women, as the women who do not cut might have a bad smell | 38 | (14.6) |
| It reduces sexual desire and regulates the sexuality of women as women have a strong sexual desire by nature | 42 | (16.1) |
| Don't know | 14 | (5.4) |
| **Performers FGC** |  |  |
| Traditional birth attendants | 60 | (27.9) |
| Doctors/nurses | 17 | (7.9) |
| Traditional circumcisers | 40 | (18.6) |
| Old women | 71 | (33.0) |
| Other | 8 | (3.7) |
| Do not know | 19 | (8.8) |
| **Who decides on performing FGC** |  |  |
| Mother | 56 | (31.6) |
| Father | 5 | (2.8) |
| Both parents | 50 | (28.3) |
| Grandmother | 29 | (16.4) |
| Religious leaders | 12 | (6.8) |
| Others | 6 | (3.4) |
| Do not know | 19 | (10.7) |
| **Is FGC common in KRI?** |  |  |
| Yes | 29 | (22.1) |
| No | 71 | (54.2) |
| Do not know | 31 | (23.7) |
| **Trends of FGC in KRI** |  |  |
| Increasing | 7 | (6.1) |
| Decreasing | 103 | (90.4) |
| Do not know | 4 | (3.5) |
| **Reasons for difference in the prevalence of FGC in different governorates** |  |  |
| Difference in the religious jurisprudence (*madhab*) | 38 | (15.7) |
| Difference in commitment to Shafi'i *madhab* | 23 | (9.5) |
| Difference in faith | 10 | (4.1) |
| Difference in people's education and awareness level | 41 | (16.9) |
| Difference in the effect of the awareness campaigns against FGC | 15 | (6.2) |
| Difference in the weather in the different regions | 30 | (12.4) |
| Tradition and cultural differences | 67 | (27.7) |
| Do not know | 18 | (7.4) |

About 59% of the religious leaders stated that people ask for their advice on FGC. More than 29% of the participants stated that men ask more, followed by educated people (17.8%) and rural people (15.4%). More than 55% of participants said that people asked about FGC more in the past; more than 61% of them stated that people complain about FGC to them, and about 58% of them stated that men complain of the loss of sexual desire of their wives. Concerning the role of religious leaders in combating FGC, more than 21% of the participants mentioned that religious leaders need to reach a conclusive answer for FGC to advise people accordingly. Around 18% of the participants said there is a need to have standard advice about FGC after carrying out adequate study and research by religious scholars and medical people. Around 16% of the participants stated that imams and preachers should have a role in banning FGC by telling the people that this is a wrong practice and is not a religious obligation. Table 3 shows the details of the attitudes of religious leaders towards FGC.

**Table 3.** Attitudes of religious leaders towards FGC.

| Variable | Frequency | Percentage |
| --- | --- | --- |
| **Do people ask for your advice on FGC** |  |  |
| Yes | 80 | (58.8) |
| No | 56 | (41.2) |
| **Frequency of asking for advice** |  |  |
| Rarely | 44 | (50.8) |
| Sometimes | 32 | (36.8) |
| Frequently | 11 | (12.6) |
| **Who asks for advice?** |  |  |
| Men | 43 | (21.3) |
| Women | 28 | (13.9) |
| Poor | 8 | (4.0) |
| Rich | 8 | (4.0) |
| Educated | 36 | (17.8) |
| Uneducated | 30 | (14.9) |
| Urban | 18 | (8.9) |
| Rural | 31 | (15.4) |
| **When did people ask more about FGC?** |  |  |
| Currently | 43 | (44.3) |
| In the past | 54 | (55.7) |
| **Were there people complaining from FGC to you?** |  |  |
| Yes | 73 | (61.3) |
| No | 46 | (38.7) |
| **Main complaints** |  |  |
| Women complain of the loss of sexual desire | 24 | (30.8) |
| Men complain of the loss of sexual desire of their wives | 45 | (57.7) |
| Other (e.g., bleeding) | 9 | (11.5) |
| **Reasons for not asking for advice** |  |  |
| There are no problems attached to FGC | 17 | (39.5) |
| People do not talk about their problems due to sensitivity | 26 | (60.5) |
| **Were people complaining of not performing FGC?** |  |  |
| Yes | 39 | (39.0) |
| No | 61 | (61.0) |
| **Role of religious leaders in combating FGC** |  |  |
| Should not be prohibited | 36 | (13.2) |
| Religious leaders could have an influential role in the issue of FGC due to the position they have in the community | 50 | (18.4) |
| The imams and preachers should have a role in banning FGC by telling people this is a wrong practice and not a religious obligation | 43 | (15.8) |
| The imams and preachers should encourage people to perform FGC | 13 | (4.8) |
| Religious leaders need to reach a conclusive answer for FGC to advise people accordingly | 58 | (21.3) |
| Imams and preachers cannot have any role in FGC issue because it is a sharia issue, and they should not intervene | 10 | (3.7) |
| Imams and preachers cannot have any role in the FGC issue because it is a very sensitive women's issue that they cannot talk about it in public during preaching | 12 | (4.4) |
| There is a need to have standard advice about FGC after carrying out adequate study and research by both religious scholars and medical people | 50 | (18.4) |

Around 14% of the participants supported performing FGC compared to 39.1% against it, and 30% who stated it should be allowed and optional. For those who support FGC, 73.9% stated it is for religious reasons compared to 26.1, which is for cultural reasons. For those against FGC, 53.5% stated that it is mainly a cultural practice with harmful effects, and 46.6% said it is due to a lack of clear religious evidence. Only 51.7% of the participants recommended banning FGC by law, and 43.5% of those who did not support banning it by law said because it will be against sharia and religious advice, followed by 37.7% who stated that such a prohibition would be mistrusted and opposed by the public. Concerning who needs to undergo FGC, 26.9% said medical professionals should decide on that. More than 77% of the participants stated that physicians or nurses should perform FGC. With regard to the religious view of FGC, the largest proportion of the participants (35.2%) stated its “honorable/*makrumah*”, while 25.9% stated it is “permissible/*mubah*”. More than 15% of the participants stated that “Islamic religion only tolerates the mildest form of FGC”.

Around 13% of participants stated “there are no clear and strong hadiths encouraging FGC” and “no hadith has banned FGC”. Half of the participants thought that the *hadith* referring to the five *fitra* (requirements of bodily hygiene) is authenticated as “*sahih*”, while 17.5% said it is related to boys only, and 14.9% said it is related to boys and girls. Concerning the *hadith* of the Prophet talking to a woman who was performing FGC in Madina, 49.1% thought it is weak (*daif*). Almost 57% of the participants considered the *hadith* that considered FGC an honorable act for women as weak (*daif*), and 80% thought that the *hadith* of “if two circumcisions meet, one should take a bath” as authenticated (*sahih*) (i.e., this text refers to a man and woman having sexual intercourse, implying that the woman as well as the man is circumcised). Table 4 shows the details of the positions of religious leaders towards FGC.

**Table 4.** Positions of religious leaders towards FGC.

| Variable | Frequency | Percentage |
| --- | --- | --- |
| **Personal position on FGC** |  |  |
| Support | 18 | (13.5) |
| Against | 52 | (39.1) |
| Allowed and be optional | 40 | (30.1) |
| No opinion | 23 | (17.3) |
| **Reasons for supporting FGC** |  |  |
| Religion | 17 | (73.9) |
| Culture | 6 | (26.1) |
| **Reasons for taking the position against FGC** |  |  |
| Lack of clear religious evidence | 27 | (46.6) |
| It is mainly a cultural practice with harmful effects | 31 | (53.5) |
| **Reasons for taking the position of allowed/optional** |  |  |
| Weak hadith which encourage FGC | 8 | (47.1) |
| No hadith prohibited the practice | 9 | (52.9) |
| **Reasons for not having opinion about position on FGC** |  |  |
| No enough information about it | 6 | (85.7) |
| Other reasons | 1 | (14.3) |
| **Would you recommend banning FGC by law?** |  |  |
| Yes | 62 | (51.7) |
| No | 58 | (48.3) |
| **Reasons for not supporting the banning of FGC** |  |  |
| Will be against sharia and religious advice | 30 | (43.5) |
| The law will not work | 13 | (18.8) |
| The law will become suspicious and people oppose it | 26 | (37.7) |
| **Who needs to undergo FGC?** |  |  |
| All girls and women | 13 | (7.0) |
| None | 28 | (15.1) |
| Those having large clitoris above the sides (labia), which makes the region ugly and over-sensitive to sexual desire, and which annoys the husband during sex | 17 | (9.1) |
| Medical professionals should decide this issue | 50 | (26.9) |
| Those living in a warm climate due to the early maturity of girls and increased sexual desire | 25 | (13.4) |
| Only for women with strong sexual desire and at risk of experiencing adultery | 19 | (10.2) |
| Do not know | 34 | (18.3) |
| **Who should perform FGC?** |  |  |
| Traditional birth attendant | 11 | (8.4) |
| Physician or nurse | 101 | (77.1) |
| People have done the job for a long time | 15 | (11.5) |
| Old women | 4 | (3.1) |
| **Religious view of FGC** |  |  |
| An obligation (*wajib*) | 8 | (7.4) |
| Sunnah | 16 | (14.8) |
| Permissible (*mubah*) | 28 | (25.9) |
| Honorable (*makrumah*) | 38 | (35.2) |
| Disliked (*makruh*) | 17 | (15.7) |
| Do not know | 1 | (0.9) |
| **Religious texts about FGC** |  |  |
| The hadiths about FGC are weak | 41 | (21.5) |
| The hadiths about FGC are inconclusive, regarding whether they refer to boy's circumcision or circumcision for boys and girls together | 12 | (6.3) |
| There are no clear and strong hadiths encouraging FGC | 25 | (13.1) |
| No hadith has banned FGC | 25 | (13.1) |
| According to available hadiths, if FGC is not encouraged, it is not prohibited, so it is allowed | 16 | (8.4) |
| In the early Islamic times, the purpose of hadiths was to regulate the FGC practice to limit the excessive cut that was practiced at that time, and did not come to encourage or obligate the practice. | 22 | (11.5) |
| Mentioning FGC in hadiths is evidence that the Prophet (peace be upon him) knew of the existence of this practice and did not prohibit it, which means it is permissible. | 21 | (11.0) |
| Islamic religion only tolerates the mildest form of FGC | 29 | (15.2) |
| **The *hadith* which refers to the five fitra** |  |  |
| Authenticated (*sahih*) | 57 | (50.0) |
| Good (*hasan*) | 10 | (8.8) |
| Weak (*daif*) | 10 | (8.8) |
| It is related to boys only | 20 | (17.5) |
| It is related to boys and girls | 17 | (14.9) |
| **The *hadith* of the Prophet talking to a woman who was performing FGC in Madina** |  |  |
| Authenticated (*sahih*) | 16 | (28.1) |
| Good (*hasan*) | 13 | (22.8) |
| Weak (*daif*) | 28 | (49.1) |
| **The *hadith* that considered FGC an honorable act for women** |  |  |
| Authenticated (*sahih*) | 12 | (20.7) |
| Good (*hasan*) | 13 | (22.4) |
| Weak (*daif*) | 33 | (56.9) |
| **The *hadith* of "If two circumcisions meet, one should take a bath"** |  |  |
| Authenticated (*sahih*) | 52 | (80.0) |
| Good (*hasan*) | 6 | (9.2) |
| Weak (*daif*) | 7 | (10.8) |

There was no statistically significant association between religious leaders’ age, education level, or work experience and their position on performing FGC. A statistically significant association (P=0.015) was found between religious leaders’ residence and their position on performing FGC. More than 46% of those residing in Duhok were against performing FGC, compared to Erbil (38.8%) and Sulaimaniyah (30%), as shown in Table 5.

**Table 5.** Association between religious leaders’ characteristics and their position on performing FGC.

| Variable | Position on performing FGC |  |  |  |  |  | Total |  | P value |
| --- | --- | --- | --- | --- | --- | --- | --- | --- | --- |
|  | Support |  | Against |  | Allowed/optional |  |  |  |  |
|  | No. | (%) | No. | (%) | No. | (%) | No. | (%) |  |
| Age groups |  |  |  |  |  |  |  |  |  |
| ≤ 40 years | 7 | (10.0) | 28 | (40.0) | 35 | (50.0) | 70 | (100) | 0.62 |
| ≥ 41 years | 11 | (17.5) | 24 | (38.1) | 28 | (44.4) | 63 | (100) |  |
| Education level |  |  |  |  |  |  |  |  |  |
| Primary/secondary | 5 | (23.8) | 4 | (19.1) | 12 | (57.1) | 21 | (100) | 0.26 |
| College/institute | 10 | (11.5) | 36 | (41.4) | 41 | (47.1) | 87 | (100) |  |
| Higher education | 4 | (16.0) | 12 | (48.0) | 9 | (36.0) | 25 | (100) |  |
| Governorate |  |  |  |  |  |  |  |  |  |
| Erbil | 11 | (12.9) | 33 | (38.8) | 41 | (48.3) | 85 | (100) | 0.015 |
| Sulaimaniyah | 7 | (35.0) | 6 | (30.0) | 7 | (35.0) | 20 | (100) |  |
| Duhok | 0 | (0.0) | 13 | (46.4) | 15 | (53.6) | 28 | (100) |  |
| Work experience (years) |  |  |  |  |  |  |  |  |  |
| ≤ 10 | 3 | (7.5) | 19 | (47.5) | 18 | (45.0) | 40 | (100) | 0.261 |
| 11 - 20 | 9 | (13.4) | 24 | (35.8) | 34 | (50.8) | 67 | (100) |  |
| ≥ 21 | 6 | (23.1) | 9 | (34.6) | 11 | (42.3) | 26 | (100) |  |

There was no statistically significant association between religious leaders’ age, education level, residence, or work experience and their position on banning FGC by law, as shown in Table 6.

**Table 6.** Association between religious leaders’ characteristics and their position on banning FGC by law.

| Variable | Banning FGC by law |  |  |  | Total |  | P value |
| --- | --- | --- | --- | --- | --- | --- | --- |
|  | Yes |  | No |  |  |  |  |
|  | No. | (%) | No. | (%) | No. | (%) |  |
| Age groups (years) |  |  |  |  |  |  |  |
| ≤ 40 | 33 | (50.8) | 32 | (49.2) | 65 | (100) | 0.831 |
| ≥ 41 | 29 | (52.7) | 26 | (47.3) | 55 | (100) |  |
| Education level |  |  |  |  |  |  |  |
| Primary/secondary | 6 | (42.9) | 8 | (57.1) | 14 | (100) | 0.603 |
| College/institute | 43 | (54.4) | 36 | (45.6) | 79 | (100) |  |
| Higher education | 10 | (45.4) | 12 | (54.6) | 22 | (100) |  |
| Governorate |  |  |  |  |  |  |  |
| Erbil | 41 | (53.9) | 35 | (46.1) | 76 | (100) | 0.152 |
| Sulaimaniyah | 7 | (33.3) | 14 | (66.7) | 21 | (100) |  |
| Duhok | 14 | (60.9) | 9 | (39.1) | 23 | (100) |  |
| Work experience (years) |  |  |  |  |  |  |  |
| ≤ 10 | 19 | (52.8) | 17 | (47.2) | 36 | (100) | 0.748 |
| 11-20 | 29 | (49.2) | 30 | (50.9) | 59 | (100) |  |
| ≥ 21 | 14 | (56.0) | 11 | (44.0) | 25 | (100) |  |

## Discussion

FGC has been practiced for centuries and it remains prevalent in many countries worldwide [19]. Three elements are considered being of great importance in the practice of FGC: religion, tradition, and patriarchy. This study shed light on religious leaders’ knowledge and attitude towards FGC and identified their position concerning FGC practice. Thus, researching such a topic among religious leaders can highlight some points that may help eradicate FGC practice in a community. One of the strengths of the current study is that it was conducted on a remarkable number of religious leaders across the KRI. Therefore, our findings could be generalizable to most of the religious leaders in the region.

FGC as a practice is usually perceived as a religious obligation. Hence, involving religious leaders in FGC campaigns to support the elimination of the practice can influence the rapid discontinuation of FGC [20,21]. Our findings indicated that the religious leaders in our study reported that they could have an influential role in the FGC issue due to their position in the community. Involvement of religious leaders in advocacy attempts against FGC in communities practicing it is quite essential [22]. Similarly, a number of religious leaders stated that they should have a role in eradicating FGC practice, as it is not a religious obligation. A study conducted in KRI also emphasized the importance of involving religious leaders in banning FGC [23]. Another study conducted by Shabila et al. [24] reported the necessity to include religious leaders in the FGC issue. Other studies concluded that the involvement of religious leaders would be critical in eradicating FGC [25,26].

However, our analysis revealed that some religious leaders highlighted the need to reach a conclusive answer concerning FGC to advise people accordingly. This is a bit worrying as it indicates that there is still no clear and concrete guidance from the religious scholars or authorized personnel with regard to the FGC issue in the KRI. Indeed, it was revealed that religious leaders believe that they need to have standard advice concerning FGC.

Our analysis showed that cultural tradition was the main reason behind practicing FGC. Previous studies likewise emphasized the role of tradition in practicing FGC elsewhere [27]. However, an earlier study conducted by Ahmed et al. [28] reported that most religious leaders considered FGC as a religious obligation. Some of them considered it as tradition merged with religion. Thus, the finding of the current study is promising in that most of the religious leaders in our sample considered FGC as a traditional practice. Holding this perspective may eventually influence people’s attitude towards FGC, as the majority of Kurdish society are Muslims and take religious leaders’ stances into great consideration. As a result, if religious leaders clarify that the FGC practice is not religiously bound to Islam [21,29], people may ultimately avoid the practice. Some well-known Kurdish Muslim scholars have argued that there is no association between Islam as a religion and FGC [17]. In other contexts, cultural traditions, controlling female sexuality, and the requirement of religion were found to be among the reasons used to justify FGC continuation among Somali communities [30].

One of the important findings of our study is that religious leaders believed that FGC is not common in KRI, and even that it is decreasing. Shabila [7] examined the trends of FGC practice between 2011-2018 in Iraq and found that the FGC prevalence decreased remarkably from 2011 to 2018 throughout all KRI governorates. Another study conducted by Koski and Heymann [31] examined the trend of FGC in 22 countries and reported that the prevalence of FGC had decreased in most regions.

Concerning the advantages and disadvantages of FGC practice, our findings highlighted that reducing or regulating women’s sexual desire to avoid sins and social problems are thought to be benefits of FGC, as well as improving cleanliness and hygiene. This is in accordance with the findings of Ahmed et al. [28], who reported similar perceptions among religious leaders. However, the disadvantages of FGC reported included reduced libido and psychological problems. There is evidence that FGC can result in various complications. Ahmed et al. [23] conducted a qualitative study exploring women’s knowledge, beliefs, and attitude about FGC and found that women expressed that FGC resulted in pain and bleeding as direct effects, and decreased sexual desire and psychological problems as long-term effects. Unsurprisingly, most women were in favor of FGC discontinuation.

It is worrying that some religious leaders still think that FGC is risk free. Health awareness activities such as seminars about the risks and disadvantages of FGC should be held in order to enlighten religious leaders at different levels of religious positions. With regard to the performers of FGC, our study revealed that religious leaders believed that old women, traditional birth attendants, and traditional circumcisers were the main performers of FGC. Healthcare professionals in KRI do not perform FGC, as reported by Shabila et al. [24]. Involving healthcare professionals and religious leaders in awareness campaigns against FGC can greatly impact eliminating FGC. Simultaneously, traditional birth attendants and traditional circumcisers should be made aware of the risks surrounding FGC and hence avoid performing it.

Our findings revealed that people, usually men, ask for religious leaders’ advice on FGC as they complain about the loss of sexual desire of their wives. Likewise, Ahmed et al. [28] reported similar complaints religious leaders encountered. Therefore, it is important to involve men in FGC eradication campaigns [32]. Women are victims of FGC, but men’s sexual lives, romantic relationships, and family dynamics can be affected by the practice. In a systematic review conducted by Varol et al. [33], it was concluded that men’s advocacy could be an important step towards the FGC elimination process. Interestingly, the study conducted by Gage et al. [26] revealed that more men illustrated “attitudinal support” towards the discontinuation of FGC.

Most religious leaders in our study were against FGC and considered it primarily to be a cultural practice. Nevertheless, some religious leaders supported the FGC practice and considered it to be a religious activity. Their beliefs on banning FGC by law were mixed, as some supported banning it while some of them did not. Another study concluded that open opposition towards FGC practice was absent among religious leaders where the practice has been banned by legislation [34]. Therefore, the Ministry of Endowment and Religious Affairs should have a clear message on this issue and make efforts to enforce the banning of FGC by law by disseminating rulings among religious leaders.

Our findings showed that the residence area is associated with the religious leaders’ position on banning FGC. For instance, in the areas where FGC is not widespread, religious leaders tend to be against the practice, compared to those areas FGC was more practiced. Again, this reinforces that FGC is practiced more based on traditional or cultural beliefs rather than because of perceived religious obligations.

## Conclusion

A conclusive decision concerning the prohibition of FGC needs to be made by religious authorities to advise people to avoid the practice. Health awareness activities incorporating FGC risks should be carried out to enlighten religious leaders at different levels of religious positions. Further research exploring perspectives of religious authorities concerning religious leaders’ inconclusive judgment about FGC is deemed necessary.

## Data Availability

All relevant data are within the manuscript and its Supporting Information files.

